# Grey and white matter volume changes after preterm birth: A meta-analytic approach

**DOI:** 10.1101/2021.06.15.21258937

**Authors:** Benita Schmitz-Koep, Bernhard Haller, Pierrick Coupé, Aurore Menegaux, Christian Gaser, Claus Zimmer, Dieter Wolke, Peter Bartmann, Christian Sorg, Dennis M. Hedderich

## Abstract

**Objective:** Lower brain grey matter volumes (GMV) and white matter volumes (WMV) have been reported at single time points in preterm-born individuals. While large MRI studies in the normative population have led to a better understanding of brain growth trajectories across the lifespan, such results remain elusive for preterm-born individuals since large, aggregated datasets of preterm-born individuals do not exist. To close this gap, we investigated GMV and WMV in preterm-born individuals as reported in the literature and contrasted it against individual volumetric data and trajectories from the general population.

**Study design:** Systematic database search of PubMed and Web of Science in March 2021 and extraction of outcome measures by two independent reviewers. Individual data on full-term controls was extracted from freely available databases. Mean GMV, WMV, total intracranial volume (TIV), and mean age at scan were the main outcome measures.

**Results:** Of 532 identified records, nine studies were included with 538 preterm-born subjects between 1.1 and 28.5 years of age. Reference data was generated from 880 full-term controls between 1 and 30 years of age. GMV was consistently lower in preterm-born individuals from infancy to early adulthood with no evidence for catch-up growth. While GMV changes followed a similar trajectory as full-term controls, WMV was particularly low in adolescence after preterm birth.

**Conclusions:** Results demonstrate altered brain volumes after premature birth across the first half of lifespan with particularly low white matter volumes in adolescence. Future studies should address this issue in large aggregated datasets of preterm-born individuals.

## INTRODUCTION

Preterm birth (PT), defined as birth <37 weeks of gestation, has a high worldwide prevalence of almost 11%.^1^ Prematurity is related to alterations in brain development in general, and in grey (GMV) and white matter volume (WMV) in particular, which has been documented at various age groups from infancy and childhood until early adulthood.^2–7^

Large population studies and open datasets have led to a better understanding of brain development and aging in the general population across the lifespan:^8–10^ Grey matter volume (GMV) rapidly increases during infancy and early childhood until it peaks at school age. A fast decrease until around 40 years of age follows with a subsequent plateau phase. Lastly, there is an accelerated decrease of GMV around 80 years of age. Development of white matter volume (WMV) follows an inverted U-shape. After fast growth in early ages, WMV peaks at around 30 to 40 years of age followed by a volume decrease. Characterization of age-dependent grey and white matter development in healthy subjects has enabled modelling pathological alterations in patients. For example, deviations from normal brain volume trajectories have been described in pathologic states such as Alzheimer’s disease and may be used as a diagnostic tool.^11^

However, structure alterations have mostly been studied in cross-sectional designs with narrow age ranges. Data from longitudinal studies with several magnetic resonance imaging (MRI) examinations are scarce and shared large datasets do not exist. Hence, aggregated evidence about brain growth trajectories of the lifespan after PT birth remains elusive.

To close this gap, we investigated GMV and WMV changes observed in cross-sectional studies of PT subjects over the first half of lifespan from infancy to early adulthood and contrasted it against brain growth trajectories of a large normative cohort of FT individuals.

## METHODS

This meta-analysis was conducted following the Preferred Reporting Items for Systematic Reviews and Meta-Analyses (PRISMA) guidelines.^12^ It was not registered. A review protocol was not prepared.

### Search strategy

The electronic databases PubMed and Web of science were systematically searched for articles published before March 30, 2021. Key words: (birth OR born) AND (preterm OR prematur*) AND (magnetic resonance imaging OR MRI) AND (brain OR intracranial) AND (volume).

### Study selection criteria

The following inclusion criteria were used to assess eligibility: (1) participants were born PT (<37 weeks of gestation); (2) MRI was used to determine mean total intracranial volume (TIV), total GMV, and total WMV; (3) mean age at MRI scan was reported and >1 year; and (4) the study was published in a peer-reviewed journal. If multiple studies reported on volumetric data of the same cohort at the same age, the study with the larger sample was included. Records were screened by two independent reviewers.

Studies with mean age lower than one year were excluded because of particular rapid brain growth in this period, in which small changes in postnatal age correspond to large volumetric changes.^13^

### Data collection process and data extraction

Data was extracted from the studies by two independent reviewers using a standardized form including author and year of publication, sample size, age at MRI scan (in years), mean and SD of TIV, GMV and WMV (in cm^3^), gestational age (GA, in weeks), birth weight (BW, in grams), percentages of male participants, year of birth, country of origin, and methodology of brain volume estimation. If PT samples were separated into groups (e.g. GA subgroups, positive or normal cranial ultrasound (cUS) findings and with or without intrauterine growth restriction (IUGR)), volumes were reported and analyzed separately. If age was reported in months, it was divided by twelve to obtain age in years. If volume was reported in mm^3^, it was divided by 1000 to obtain volume in cm^3^. If volume was reported in dm^3^, it was multiplied with 1000 to obtain volume in cm^3^. One study^7^ reported mean GMV/TIV-ratio and mean WMV/TIV ratio. Mean GMV and WMV values were calculated from these ratios. If BW was reported in kilograms, it was divided by 1000 to obtain weight in grams.

### Data on full-term controls

Data on FT controls was extracted from freely available databases: C-MIND (https://research.cchmc.org/c-mind/), NDAR-NIHPD (http://www.bic.mni.mcgill.ca/nihpd/info/data_access.html), ICBM (http://www.loni.usc.edu/ICBM/), and IXI (http://brain-development.org/ixi-dataset/). TIV, GMV and WMV were calculated from these four datasets as previously described by Coupé et al.^10^ Overall, 880 FT controls between 0.7 and 30 years of age were included: 236 participants from C-MIND (mean age=8.4 years, age range=0.7–18.9 years, male=45.3%), 375 from NDAR-NIHPD (mean age=11.9 years, age range=1.1–29.0 years, male=53.6%), 169 from ICBM (mean age=23.9 years, age range=18–30 years, male=52.7%), and 100 from IXI (mean age=25.7 years, age range=20.0–29.8 years, male=44%).

### Statistical analysis

Statistical analysis was performed with R (R Foundation for Statistical Computing, 2020, Vienna, Austria).^14^ A nonlinear regression model (Model 6, the cubic hybrid model, from Coupé et al.:^10^ 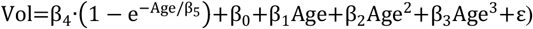 was used to model GMV, WMV, TIV, GMW/TIV, and WMV/TIV for FT controls in dependence of age using the R library *minpack*.*lm*.^15^ Grey areas surrounding the curves show 95%-confidence intervals (CI), dotted lines show 95%-prediction intervals (PI), which were calculated using the R library *propagate*.^16^ Mean values extracted for PT subjects from articles are included as diamonds in Figures 2 and 3. The size of each diamond is proportional to the sample size of the study.

## RESULTS

### Study selection and characteristics

The search strategy identified 532 records (Figure 1, Table 1). After screening of abstract and title, 167 articles were identified as potentially relevant. Based on the inclusion criteria, nine studies were eligible for the meta-analysis.^5,7,17–23^ Most of the studies excluded did not report on all variables of interest, i.e., TIV, GMV and WMV. For example, a recent study tracking regional brain growth up to age 13 in children born term and very preterm appeared to meet the inclusion criteria. However, while it did report mean age at scan, mean TIV and mean WMV, it did not report mean global GMV.^24^ Therefore, it was excluded. Another study investigating regional brain volume abnormalities and long-term cognitive outcome in preterm infants reported adjusted marginal mean of specific brain regions, however, it did not report mean TIV, WMV or GMV.^25^ Therefore, it was excluded. Seven studies were excluded because they reported on data from cohorts that were already covered by other studies. In summary, 538 PT subjects collected in studies with mean age between 1.1 and 28.5 years, and data from 880 FT controls between 0.7 and 30 years of age were analyzed. Please see Figure 1 for a flowchart depicting study selection. Study characteristics of the nine studies included are shown in Table 1.

**Table 1:** Characteristics of preterm studies included.

| Author (year) | Group | Sample (n) | Age at scan (years) | TIV mean (cm<sup>3</sup>) | TIV SD (cm<sup>3</sup>) | GMV mean (cm<sup>3</sup>) | GMV SD (cm<sup>3</sup>) | WMV mean (cm<sup>3</sup>) | WMV SD (cm<sup>3</sup>) | GA (weeks) | BW (g) | Male (%) | Year of birth | Country of origin | Methodology of brain volume estimation |
| --- | --- | --- | --- | --- | --- | --- | --- | --- | --- | --- | --- | --- | --- | --- | --- |
| Pascoe (2019)<sup>17</sup> |  | 150 | 28.5 | 1504 | 140 | 669 | 61 | 513 | 61 | 28.8 | 1077 | 41.3 | 1986 | New Zealand | CAT12 toolbox (SPM12) |
| Lemola (2017)<sup>18</sup> |  | 57 | 10.0 | 1397 | 148 | 777 | 70 | 464 | 67 | 29.7 | 1447 | 65.3 | 1998-2006 | Switzerland | New segment toolbox (SPM8) |
| Meng (2016)<sup>7</sup> |  | 85 | 26.5 | 1385 |  | 609.4 |  | 554 |  | 30.67 | 1356 | 55.3 | 1985-1986 | Germany | VBM8 toolbox (SPM8) |
| Northam (2011)<sup>19</sup> | positive cUS | 27 | 16 | 1601 | 222 | 757 | 57 | 418 | 46 | 27.1 | 1081 | 44.4 | 1989-1994 | United Kingdom | VBM5 toolbox (SPM5) |
| Northam (2011)<sup>19</sup> | normal cUS | 22 | 16.3 | 1480 | 193 | 765 | 65 | 438 | 49 | 28.1 | 1098 | 31.8 | 1989-1994 | United Kingdom | VBM5 toolbox (SPM5) |
| Padilla (2011)<sup>20</sup> | IUGR | 18 | 1.1* | 969.6 | 101.8 | 683.7 | 64.5 | 243.5 | 36.9 | 32.1 | 1060 | 38.9 | 2006-2007 | Spain | VBM5 toolbox (SPM5) |
| Padilla (2011)<sup>20</sup> | AGA | 15 | 1.1* | 1001.1 | 95.4 | 714.0 | 57.0 | 240.7 | 38.0 | 31 | 1580 | 73.3 | 2006-2007 | Spain | VBM5 toolbox (SPM5) |
| Soria-Pastor (2009)<sup>5</sup> |  | 20 | 9.3 | 1641.2 | 172.6 | 821.7 | 84.9 | 419.2 | 53.8 | 32.5 | 1754 | 55 | 1996-1998 | Spain | SPM5 |
| Narberhaus (2007)<sup>21</sup> | GA ≤ 27 | 9 | 14.1 | 1354.8 | 174.1 | 733.4 | 54.7 | 312.1 | 57.9 | 26.4 | 899 | 77.8 | 1983-1994 | Spain | SPM2 |
| Narberhaus (2007)<sup>21</sup> | GA 28 - 30 | 19 | 14.6 | 1488.5 | 164.9 | 771.3 | 133.2 | 377.2 | 57.9 | 29 | 1140 | 42.1 | 1983-1994 | Spain | SPM2 |
| Narberhaus (2007)<sup>21</sup> | GA 31 - 33 | 25 | 13.8 | 1445.3 | 146.4 | 778.1 | 72.1 | 372.0 | 52.1 | 31.7 | 1534 | 44 | 1983-1994 | Spain | SPM2 |
| Narberhaus (2007)<sup>21</sup> | GA 34 - 36 | 11 | 13.55 | 1473.3 | 148.4 | 780.3 | 69.9 | 389.8 | 45.1 | 34.6 | 2446 | 63.6 | 1983-1994 | Spain | SPM2 |
| Gimenez (2006b)<sup>23</sup> |  | 50 | 14.5 | 1488.8 | 148.8 | 787.8 | 80.8 | 377.2 | 47.4 | 29.9 | 1327 | 48 | 1982-1994 | Spain | SPM2 |
| Gimenez (2006a)<sup>22</sup> |  | 30 | 14.3 | 1460 | 140 | 780 | 70 | 360 | 50 | 29.1 | 1108 | n.a. | n.a. | Spain | SPM2 |

Abbreviations: AGA, appropriate for gestational age; BW, birth weight; cUS, cranial ultrasound; GA, gestational age; GMV, grey matter volume; IUGR, intrauterine growth
restriction; SD, standard deviation; TIV, total intracranial volume; WMV, white matter volume
\* corrected age

**Figure 1:**
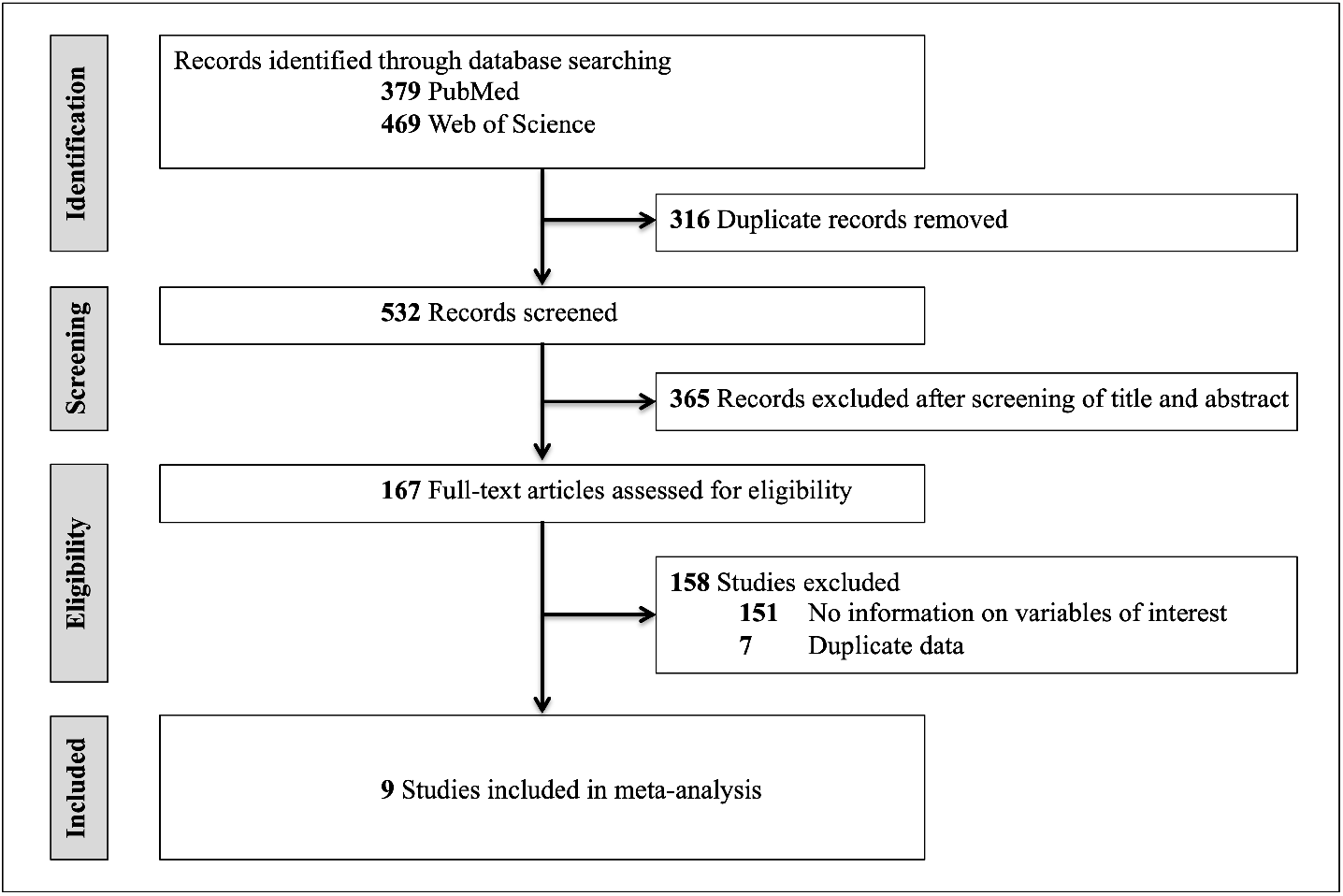
Flowchart of study selection.

### Development of grey matter volume after preterm birth

Figure 2a illustrates mean absolute GMV extracted from the PT studies contrasted against individual GMV data of FT controls. Mean absolute GMV in all of the PT studies was below mean GMV in FT development. One study^20^ reported mean absolute GMV of PT infants appropriate for gestational age (AGA) at about 1 year of age just within the 95%-CI. All other studies reported mean absolute GMV below the 95%-CI. The highest mean absolute GMV was reported at school age. While mean absolute GMV decreased between adolescence and early adulthood in both PT subjects and FT controls, this decrease appeared steeper in the PT studies.

Figure 3a illustrates GMV/TIV, i.e., relative GMV, from the PT studies contrasted against individual data of FT controls. Relative GMV decreased from infancy through early adulthood. Relative GMV calculated from data on infants with and without IUGR reported by Padilla et al.^20^ was above the curve describing FT development. All other values were below the 95%-CI. Data from three PT studies of school-aged children^5^, adolescents with brain injury^19^ and adults^17^ were below the 95%-PI.

**Figure 2:**
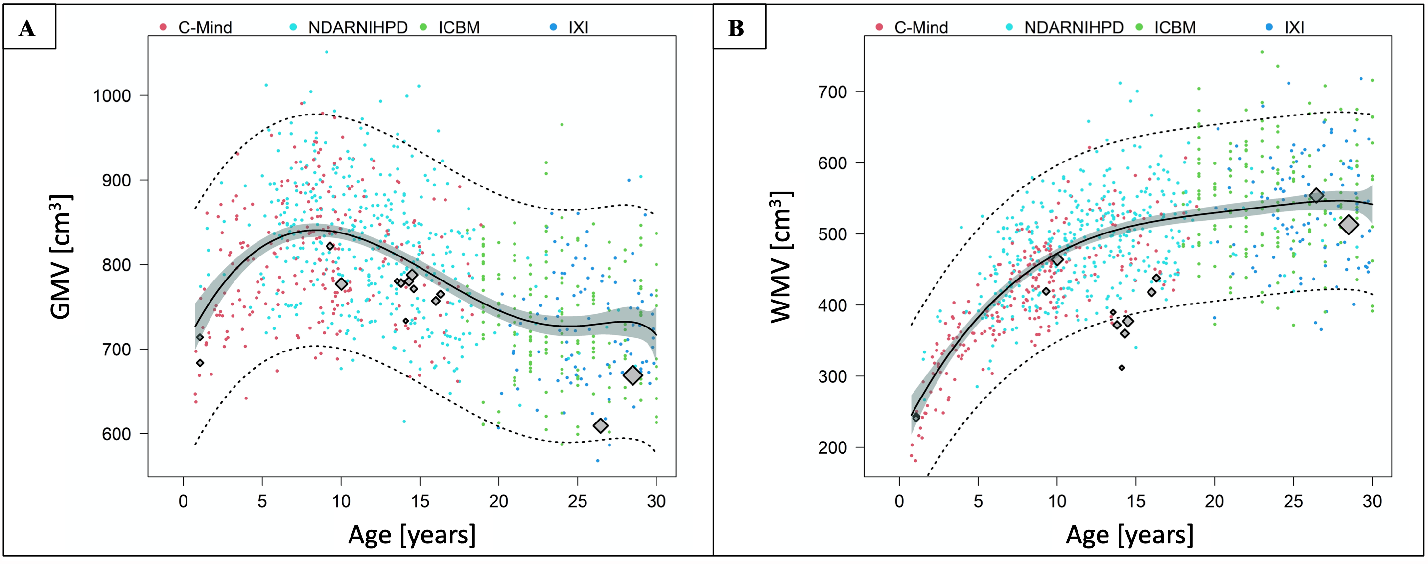
**A. Mean absolute grey matter volume after preterm birth compared to full-term controls**. The graph shows mean absolute GMV extracted from the PT studies contrasted against the GMV trajectory of FT controls with 95%-CI (grey areas) and 95%-PI (dotted lines). **B. Mean absolute white matter volume after preterm birth compared to full-term controls**. The graph shows mean absolute WMV extracted from the PT studies contrasted against the WMV trajectory of FT controls with 95%-CI (grey areas) and 95%-PI (dotted lines). The four different datasets on FT controls are illustrated in different colors. C-MIND is presented in red, NDAR-NIHPD in in cyan, ICBM in green, and IXI in blue. Mean volumes from the PT studies are included as diamonds. The size of each diamond is proportional to the sample size of the study. Abbreviations: CI, confidence interval; FT, full-term; GMV, grey matter volume; PI, prediction interval; PT, preterm; WMV, white matter volume.

**Figure 3:**
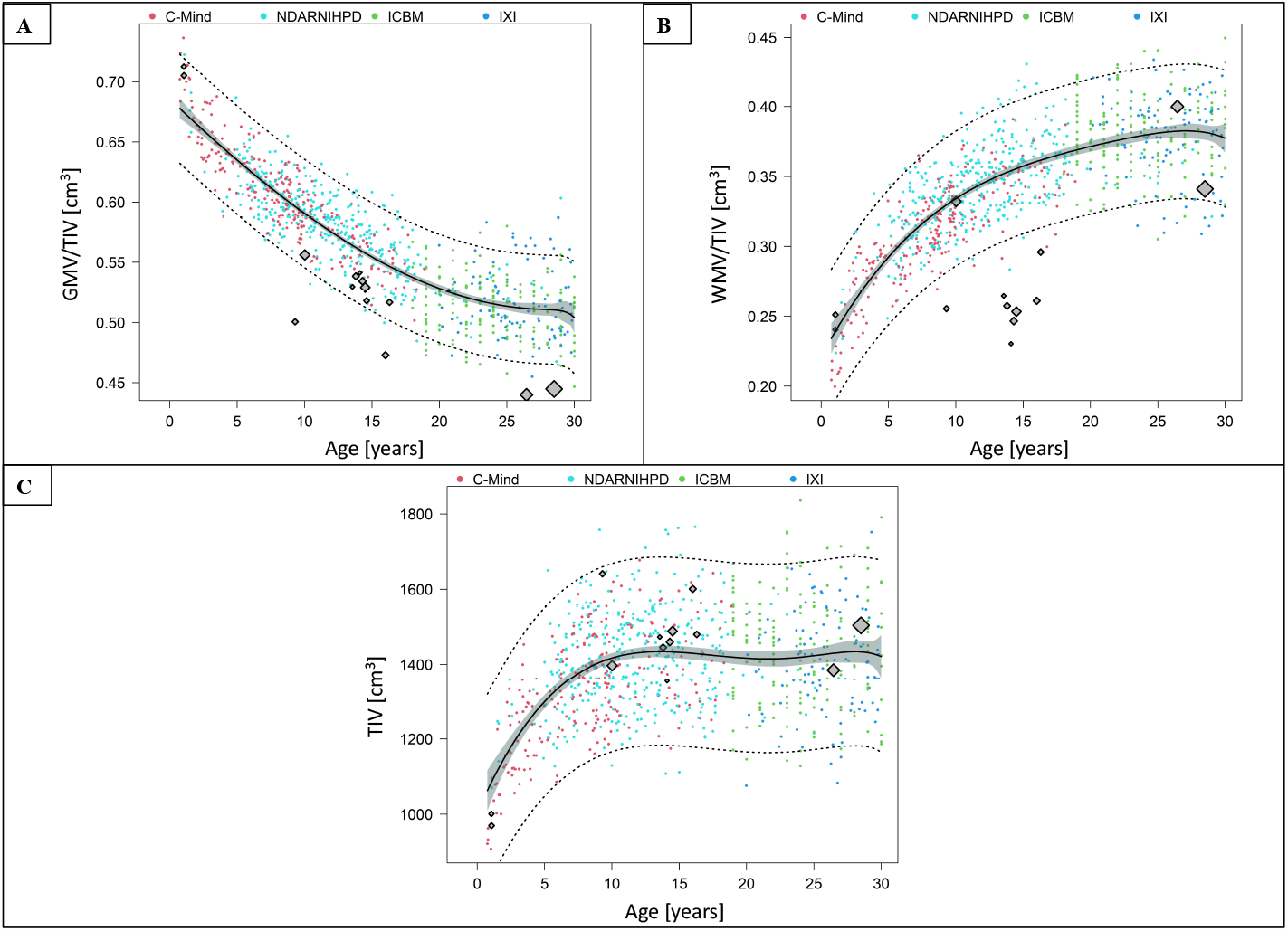
**A. Relative grey matter volume after preterm birth compared to full-term controls**. The graph shows GMV/TIV extracted from the PT studies contrasted against the GMV/TIV trajectory of FT controls with 95%-CI (grey areas) and 95%-PI (dotted lines). **B. Relative white matter volume after preterm birth compared to full-term controls**. The graph shows WMV/TIV extracted from the PT studies contrasted against the WMV/TIV trajectory of FT controls with 95%-CI (grey areas) and 95%-PI (dotted lines). **C. Total intracranial volume after preterm birth compared to full-term controls**. The graph shows mean TIV extracted from the PT studies contrasted against the TIV trajectory of FT controls with 95%-CI (grey areas) and 95%-PI (dotted lines). The four different datasets on FT controls are illustrated in different colors. C-MIND is presented in red, NDAR-NIHPD in in cyan, ICBM in green, and IXI in blue. Data points from the PT studies are included as diamonds. The size of each diamond is proportional to the sample size of the study. Abbreviations: CI, confidence interval; FT, full-term; GMV, grey matter volume; PI, prediction interval; PT, preterm; TIV, total intracranial volume; WMV, white matter volume.

In summary, GMV was lower after PT birth from infancy through early adulthood while following a similar trajectory as FT controls.

### Development of white matter volume after preterm birth

Figure 2b illustrates mean absolute WMV extracted from the PT studies contrasted against individual WMV data of FT controls. Except for one study in early adulthood^7^, mean absolute WMV in all of the PT studies was below the curve describing mean FT development. Mean absolute WMV of infants with and without IUGR reported by Padilla et al.^20^ was within the 95%-CI. All other studies reported mean absolute WMV below the 95%-CI. Mean absolute WMV reported in adolescence was particularly low, moving away from the curve describing mean FT development.

Figure 3b illustrates WMV/TIV, i.e., relative WMV, from the PT studies contrasted against individual data of FT controls. Relative WMV calculated from data on infants with and without IUGR reported by Padilla et al.^20^ and from data in early adulthood^7^ was above the curve describing FT development. Data from one PT study at school age^18^ was below the curve describing FT development but just within the 95%-CI. All other values were below the curve. The other PT study at school age^5^ and all PT studies in adolescents were below the 95%-PI.

In summary, we observed particularly low WMV in adolescence, while WMV in infancy and early adulthood were closer to the trajectory describing FT development.

### Total intracranial volume after preterm birth

Figure 3c illustrates mean TIV extracted from the PT studies contrasted against individual TIV data of FT controls. While TIV reported by some studies in infancy, childhood and early adulthood was below the curve describing FT development, other PT studies reported higher values

## DISCUSSION

This meta-analysis investigated changes of GMV and WMV after preterm birth compared to full-term controls across the first half of lifespan. We found lower GMV after preterm birth from infancy until adulthood, indicating lastingly altered brain development. GMV changes of preterm-born individuals followed a similar trajectory as full-term controls with no evidence for catch-up growth. WMV was particularly low in adolescence after preterm birth. Furthermore, the present study highlights the need for large longitudinal datasets to compare preterm and full-term brain development across the lifespan.

### Changes of grey and white matter volume after preterm birth across the first half of lifespan

We found lower GMV after PT birth compared to FT controls from infancy through early adulthood. Similar to their FT peers, GMV in PT subjects peaked at school age and decreased between adolescence and early adulthood. However, relative GMV in infancy was higher compared to FT peers, possibly reflecting delayed maturation. Yet this speculation is based on only one study. At school age, in adolescence and in early adulthood GMV was lower. There was no evidence for catch-up growth. Furthermore, WMV after PT birth was particularly low in adolescence.

Comparing our results to the scarce longitudinal data available is difficult because of methodological differences. Ment et al.^26^ observed reduction in cerebral GMV and an increase in WMV between 8 and 12 years of age. However, compared to FT controls, there was lower GMV reduction and less WMV gain over time^26^. In contrast, our results show that GMV in PT individuals mostly follow the FT reference curve, albeit on an overall lower level and that WMV is particularly low in early adolescence. Mostly in line with our results, Parker et al.^27^ reported significantly smaller GMV and WMV in VP subjects compared to FT controls in adolescence (15 years) and in early adulthood (19 years). Furthermore, they reported similar growth patterns between these two timepoints^27^. Karolis et al.^28^ observed GMV alterations in VP subjects between adolescence (15 years), early adulthood (20 years) and adulthood (30 years), indicating accelerated brain maturation. Similarly, our findings of lower relative GMV at school age, in adolescence and early adulthood might indicate early aging. Consistent with our findings, Karolis et al.^28^ found no evidence for developmental catch-up. In line with our results, de Kieviet et al.^29^ found reductions in GMV and WMV compared to FT controls in their investigation of brain volume throughout childhood and adolescence in subjects born VP and with very low birthweight in a meta-analysis.

Remarkably, our results suggest a significant impact of prematurity on WMV in adolescence. On a cellular level, pre-oligodendrocytes, precursors of mature oligodendrocytes, are critical for WM myelination. Pre-oligodendrocytes are specifically vulnerable to insults, such as ischemia and inflammation, resulting in cell injury or death and subsequent replenishment but dysmaturation.^6,30–32^ Hence, our results showing preferential alteration of WMV might emphasize the significance of pre-oligodendrocyte vulnerability and dysmaturation in the context of prematurity.

In conclusion, PT birth has lasting effects on the development of GMV and WMV compared to FT controls with particularly low WMV in adolescence. We found no evidence for GMV catch-up growth.

### What is needed in the future?

PT cohorts have been followed from birth and well investigated, however, to date longitudinal imaging data on PT subjects are scarce and only cover specific age ranges. Shared large datasets do not exist. Methodological differences in image processing as well as statistical analysis make it difficult to compare results. Thanks to large, open datasets on healthy subjects, FT developmental trajectories have been well characterized,^10^ facilitating insights in diseases affecting brain structure, for example Alzheimer’s Disease.^11^ To achieve similar goals for PT developmental trajectories, access to original imaging data of large PT populations across all age groups as well as shared individual patient data with information on parameters like birthweight, duration of neonatal intensive care unit exposure, and days of mechanical ventilation, and information about possible brain damage, such as intraventricular hemorrhage, is critical.

### Limitations

There are limitations of this meta-analysis. First, only data on limited age groups was available that fit the inclusion criteria since raw volumes were rarely reported. Hence, PT trajectories of GMV and WMV in comparison to FT trajectories remain less clear in some age groups, for example between about 2 and 7 years. However, it is inherently difficult to obtain a brain MRI in this age group since this is generally not possible without general anesthesia.

Second, as mentioned above, different methods of processing images make it very difficult to compare data. All PT studies included in this meta-analysis used toolboxes within SPM (https://www.fil.ion.ucl.ac.uk/spm/) for segmentation, while FT images were processed with volBrain (https://www.volbrain.upv.es).^33^ Hence, the possibility of systematic error cannot be ruled out. Only access to raw imaging data could improve this major limitation and, as discussed, is needed. However, contrasting individual MRI scans and derived measures of GMV and WMV to reference data from open datasets is increasingly used in clinical routine. Lastly, GMV and WMV are global measures representing brain development. However, determination of developmental changes of other measures such as gyrification and individual volumes of subcortical structures are necessary to characterize PT brain development, but data is not available.

## CONCLUSION

In conclusion, GMV is lower after PT birth from infancy through early adulthood, indicating lastingly altered brain development. GMV changes were similar to the trajectory of full-term controls with no evidence for catch-up growth. WMV was particularly low in adolescence after preterm birth. Large longitudinal datasets are crucial to compare PT with FT brain development across the lifespan.

## Supporting information

PRISMA

## Data Availability

Data from published articles.

## ABBREVIATIONS

AGA: appropriate for gestational age
BW: birth weight
CI: confidence interval
cUS: cranial ultrasound
FT: full-term
GA: gestational age
GMV: grey matter volume
IUGR: intrauterine growth restriction
MRI: magnetic resonance imaging
PT: preterm
SD: standard deviation
SPM: Statistical Parametric Mapping
TIV: total intracranial volume
VP: very preterm
WMV: white matter volume

## ACKNOWLEDGEMENTS

We would like to thank all investigators involved in collecting following datasets: C-MIND (https://research.cchmc.org/c-mind/), NDAR-NIHPD (http://www.bic.mni.mcgill.ca/nihpd/info/data_access.html), ICBM (http://www.loni.usc.edu/ICBM/), and IXI (http://brain-development.org/ixi-dataset/).

This work was supported by the Deutsche Forschungsgemeinschaft (SO 1336/1-1 to C.S.), German Federal Ministry of Education and Science (BMBF 01ER0801 to P.B. and D.W., BMBF 01ER0803 to C.S.), the RECAP preterm project, an EU Horizon 2020 study (supported by grant 733280; D.W. and P.B.), and the Kommission für Klinische Forschung, Technische Universität München (KKF 8765162 to C.S., KKF 8700000474 to D.M.H., and KKF 8700000620 to B.S.-K.).

## REFERENCES

1. Chawanpaiboon S, Vogel JP, Moller A-B, et al. Global, regional, and national estimates of levels of preterm birth in 2014: a systematic review and modelling analysis. Lancet Glob Heal. 2019;7(1):e37–e46. doi:10.1016/S2214-109X(18)30451-0

2. Nosarti C, Al-Asady MHS, Frangou S, Stewart AL, Rifkin L, Murray RM. Adolescents who were born very preterm have decreased brain volumes. Brain. 2002;125(Pt 7):1616–1623. doi:10.1093/brain/awf157

3. Nosarti C, Nam K-W, Walshe M, et al. Preterm birth and structural brain alterations in early adulthood. NeuroImage Clin. 2014;6(C):180–191. doi:10.1016/j.nicl.2014.08.005

4. Inder TE, Warfield SK, Wang H, Hüppi PS, Volpe JJ. Abnormal cerebral structure is present at term in premature infants. Pediatrics. 2005;115(2):286–294. doi:10.1542/peds.2004-0326

5. Soria-Pastor S, Padilla N, Zubiaurre-Elorza L, et al. Decreased Regional Brain Volume and Cognitive Impairment in Preterm Children at Low Risk. Pediatrics. 2009;124(6):e1161–e1170. doi:10.1542/peds.2009-0244

6. Volpe JJ. Brain injury in premature infants: a complex amalgam of destructive and developmental disturbances. Lancet Neurol. 2009;8(1):110–124. doi:10.1016/S1474-4422(08)70294-1

7. Meng C, Bäuml JG, Daamen M, et al. Extensive and interrelated subcortical white and gray matter alterations in preterm-born adults. Brain Struct Funct. 2016;221(4):2109– 2121. doi:10.1007/s00429-015-1032-9

8. Hedman AM, van Haren NEM, Schnack HG, Kahn RS, Hulshoff Pol HE. Human brain changes across the life span: a review of 56 longitudinal magnetic resonance imaging studies. Hum Brain Mapp. 2012;33(8):1987–2002. doi:10.1002/hbm.21334

9. Giedd JN, Blumenthal J, Jeffries NO, et al. Brain development during childhood and adolescence: a longitudinal MRI study. Nat Neurosci. 1999;2(10):861–863. doi:10.1038/13158

10. Coupé P, Catheline G, Lanuza E, Manjón JV, Alzheimer’s Disease Neuroimaging Initiative. Towards a unified analysis of brain maturation and aging across the entire lifespan: A MRI analysis. Hum Brain Mapp. 2017;38(11):5501–5518. doi:10.1002/hbm.23743

11. Coupé P, Manjón JV, Lanuza E, Catheline G. Lifespan Changes of the Human Brain In Alzheimer’s Disease. Sci Rep. 2019;9(1):3998. doi:10.1038/s41598-019-39809-8

12. Moher D, Liberati A, Tetzlaff J, et al. Preferred reporting items for systematic reviews and meta-analyses: The PRISMA statement. PLoS Med. 2009;6(7). doi:10.1371/journal.pmed.1000097

13. Dobbing J, Sands J. Quantitative growth and development of human brain. Arch Dis Child. 1973;48(10):757–767. doi:10.1136/adc.48.10.757

14. R Core Team. R: A language and environment for statistical computing. 2020.

15. Elzhov T V, Mullen KM, Spiess A-N, Bolker B. minpack.lm: R Interface to the Levenberg-Marquardt Nonlinear Least-Squares Algorithm Found in MINPACK, Plus Support for Bounds. 2016. https://cran.r-project.org/web/packages/minpack.lm/minpack.lm.pdf.

16. Spiess A-N. propagate: Propagation of Uncertainty. 2018. https://cran.r-project.org/package=propagate.

17. Pascoe MJ, Melzer TR, Horwood LJ, Woodward LJ, Darlow BA. Altered grey matter volume, perfusion and white matter integrity in very low birthweight adults. NeuroImage Clin. 2019;22:101780. doi:10.1016/j.nicl.2019.101780

18. Lemola S, Oser N, Urfer-Maurer N, et al. Effects of gestational age on brain volume and cognitive functions in generally healthy very preterm born children during school-age: A voxel-based morphometry study. Lidzba K, ed. PLoS One. 2017;12(8):e0183519. doi:10.1371/journal.pone.0183519

19. Northam GB, Liégeois F, Chong WK, S. Wyatt J, Baldeweg T. Total brain white matter is a major determinant of IQ in adolescents born preterm. Ann Neurol. 2011;69(4):702–711. doi:10.1002/ana.22263

20. Padilla N, Falcón C, Sanz-Cortés M, et al. Differential effects of intrauterine growth restriction on brain structure and development in preterm infants: A magnetic resonance imaging study. Brain Res. 2011;1382:98–108. doi:10.1016/j.brainres.2011.01.032

21. Narberhaus A, Segarra D, Caldú X, et al. Gestational Age at Preterm Birth in Relation to Corpus Callosum and General Cognitive Outcome in Adolescents. J Child Neurol. 2007;22(6):761–765. doi:10.1177/0883073807304006

22. Giménez M, Junqué C, Narberhaus A, Botet F, Bargalló N, Mercader JM. Correlations of thalamic reductions with verbal fluency impairment in those born prematurely. Neuroreport. 2006;17(5):463–466. doi:10.1097/01.wnr.0000209008.93846.24

23. Giménez M, Junqué C, Narberhaus A, Bargalló N, Botet F, Mercader JM. White matter volume and concentration reductions in adolescents with history of very preterm birth: A voxel-based morphometry study. Neuroimage. 2006;32(4):1485–1498. doi:10.1016/j.neuroimage.2006.05.013

24. Thompson DK, Matthews LG, Alexander B, et al. Tracking regional brain growth up to age 13 in children born term and very preterm. Nat Commun. 2020;11(1):696. doi:10.1038/s41467-020-14334-9

25. Peterson BS, Vohr B, Staib LH, et al. Regional brain volume abnormalities and longterm cognitive outcome in preterm infants. J Am Med Assoc. 2000;284(15):1939–1947. doi:10.1001/jama.284.15.1939

26. Ment LR, Kesler S, Vohr B, et al. Longitudinal brain volume changes in preterm and term control subjects during late childhood and adolescence. Pediatrics. 2009;123(2):503–511. doi:10.1542/peds.2008-0025

27. Parker J, Mitchell A, Kalpakidou A, et al. Cerebellar growth and behavioural & neuropsychological outcome in preterm adolescents. Brain. 2008;131(Pt 5):1344–1351. doi:10.1093/brain/awn062

28. Karolis VR, Froudist-Walsh S, Kroll J, et al. Volumetric grey matter alterations in adolescents and adults born very preterm suggest accelerated brain maturation. Neuroimage. 2017;163(September):379–389. doi:10.1016/j.neuroimage.2017.09.039

29. de Kieviet JF, Zoetebier L, van Elburg RM, Vermeulen RJ, Oosterlaan J. Brain development of very preterm and very low-birthweight children in childhood and adolescence: a meta-analysis. Dev Med Child Neurol. 2012;54(4):313–323. doi:10.1111/j.1469-8749.2011.04216.x

30. Volpe JJ. Dysmaturation of Premature Brain: Importance, Cellular Mechanisms, and Potential Interventions. Pediatr Neurol. 2019;95:42–66. doi:10.1016/j.pediatrneurol.2019.02.016

31. Back SA, Han BH, Luo NL, et al. Selective vulnerability of late oligodendrocyte progenitors to hypoxia-ischemia. J Neurosci. 2002;22(2):455–463. doi:10.1523/JNEUROSCI.22-02-00455.2002

32. Segovia KN, McClure M, Moravec M, et al. Arrested oligodendrocyte lineage maturation in chronic perinatal white matter injury. Ann Neurol. 2008;63(4):520–530. doi:10.1002/ana.21359

33. Manjón J V., Coupé P. volBrain: An Online MRI Brain Volumetry System. Front Neuroinform. 2016;10(JUL):1–14. doi:10.3389/fninf.2016.00030

